# Maternal synapsin 1 autoantibodies are associated with neurodevelopmental delay

**DOI:** 10.1101/2022.11.17.22282130

**Authors:** Isabel Bünger, Konstantin L. Makridis, Jakob Kreye, Marc Nikolaus, Eva Sedlin, Tim Ullrich, Helle Foverskov Rasmussen, Dragomir Milovanovic, Christian Hoffmann, Johannes Tromm, Markus Höltje, Harald Prüss, Angela M. Kaindl

**Author notes:** **Corresponding author:** Prof. Dr. med Angela M. Kaindl, Pediatric Neurology, Charité – Universitätsmedizin Berlin, Augustenburger Platz 1, 13353 Berlin, Germany. These authors contributed equally to this work and share first authorship (Isabel Bünger, Konstantin L. Makridis). These authors contributed equally to this work and share last authorship (Harald Prüss, Angela M. Kaindl).

## Abstract

Maternal autoantibodies can be transmitted diaplacentally, with potentially deleterious effects on neurodevelopment. Synapsin 1 (SYN1) is a neuronal protein that is important for synaptic communication and neuronal plasticity. While monoallelic loss of function (LoF) variants in the *SYN1* gene result in X-linked intellectual disability (ID), learning disabilities, epilepsy, behavioral problems, and macrocephaly, the effect of SYN1 autoantibodies on neurodevelopment remains unclear. We recruited a clinical cohort of 208 mothers and their children with neurologic abnormalities and analyzed the role of maternal SYN1 autoantibodies. We identified seropositivity in 9.6% of mothers, and seropositivity was associated with an increased risk for ID and behavioral problems. Furthermore, children more frequently had epilepsy, macrocephaly, and developmental delay, in line with the SYN1 LoF phenotype. Whether SYN1 autoantibodies have a direct pathogenic effect on neurodevelopment or serve as biomarkers requires functional experiments.

## Introduction

Synapsin 1 (SYN1) is a neuronal phosphoprotein encoded by the *SYN1* gene and plays an important role in neurotransmitter release and neuronal plasticity (1, 2). Patients with loss of function (LoF) variants in the *SYN1* gene have been associated with X-linked phenotypes consisting of epilepsy, learning difficulties, intellectual disability (ID), macrocephaly, behavioral problems, and autism-spectrum disorders (ASD) (MIM#300491, MIM#300115) (3-5).

In recent years, anti-neuronal autoantibodies have been increasingly described to cause target-specific autoimmune neurologic disorders, in some cases matching the clinical phenotypes of patient carrying genetic variants of these targets. For example, both patients with variants in GABA_A_ receptor subunit genes and those with GABA_A_ receptor autoantibodies display epilepsy as a common phenotype (6-8). Autoantibodies are usually regarded as those generated within an individual; however, they can also be transferred diaplacentally from mother to child (9). As the blood brain barrier is not yet fully matured, these autoantibodies of maternal origin can target the developing brain and thereby potentially affect neurodevelopment (9). There is increasing evidence that maternal immune activation through autoantibodies can influence the occurrence of neurodevelopmental disorders, such as ASD (9). In addition, multiple murine models have demonstrated the direct deleterious effect of various anti-neuronal autoantibodies in offspring through gestational transfer (10-13).

In previous studies, autoantibodies against SYN1 have been identified in patients with neurologic and psychiatric disorders (14, 15). We have recently shown that SYN1 autoantibodies in pregnant women are associated with abnormalities of fetal development including intrauterine growth retardation (Preprint) (16). However, their role in mothers of children with defined neurologic disorders has not been studied so far. Thus, we assessed the prevalence of SYN1 autoantibodies in mothers of a cohort of 208 pediatric patients with neurological disorders.

## Material and Methods

### Cell-based assay

Synapsin 1b (SYN1b) cell-based assay (CBA) was performed using human embryonic kidney (HEK293T) cells which were transfected with human SYN1b. Methanol-fixed cells were then incubated with sera diluted 1:300. IgG AF488-antibody (Dianova, #109-545-003), commercial rabbit SYN1/SYN2 antibody (Synaptic Systems, #106002) and anti-rabbit IgG AF594-antibody (Jackson IR, #111-585-003) were used to detect IgG binding. Two independent investigators scored the CBA using the following semi-quantitative score: 0, no binding; 1, unspecific signal (‘background’); 2, positive (intensive binding). Sera were tested on control cells overexpressing the NR1 subunit of the N-methyl-D-aspartate receptor (NMDAR), contactin-associated protein 2 (Caspr2), and SYN1 CBA-positive sera also on untransfected control HEK293T cells to exclude non-specific binding (14, 16).

### Enzyme-linked immunosorbent assay

For the ELISA, 200 ng of recombinant rat Syn1-His-EGFP (17) was diluted in PBS and incubated overnight at 4 °C in 96-well high-binding plates. Serum (1:200 diluted in PBS containing 1% BSA, 0.05% Tween) was incubated for one hour at room temperature, on which HRP-coupled anti-human IgG antibody (Dianova, #109-035-003) was added for an additional hour. Ultra TMB substrate solution was added, and reaction terminated after one minute with H_2_SO_4_. Absorbance at 450 nm was measured and corrected with controls (mean of wells without serum and reference absorbance at 630 nm).

### Western blot

SDS-Page and Western blotting were performed as described using cortices from *Syn1/2/3* triple knockout (TKO) mice and wild type (WT) mice (14). Sera of CBA with scores of 1-2 were diluted to 1:200 and used to incubate membranes. A rabbit polyclonal SYN1/2 antibody (#106002) was used as positive control and a mouse monoclonal antibody against glyceraldehyde-3-phosphate dehydrogenase (GAPDH, Merck Millipore, #MAB374) served as loading control.

### Clinical data and statistical analysis

We studied a cohort of 208 mothers of pediatric patients with neurological disorders treated at the Center for Chronically Sick Children, Charité – Universitätsmedizin Berlin, Germany. Patient data were collected blinded to maternal antibody levels using a standardized data collection sheet. Statistical analysis was performed using R (RStudio version 4.2.1) and the packages *crosstable, UpSetR*, and *psych*. Descriptive statistics were performed to calculate percentages and frequencies. Differences in categorical variables are reported as odds ratio and 95% confidence interval. P values were calculated using the Chi-square test and Fisher’s Exact Probability Test, as applicable. Test results with p<0.05 were considered statistically significant. The study was approved by the local ethics committee (approval no. #EA2/220/20).

## Results

We screened sera for SYN1 autoantibodies in 208 mothers to assess the relationship with neurodevelopment in children. Due to the explorative approach, no exclusion criteria were set. Mothers gave birth at an average age of 30.93 ±5.81 years. Children (male n=134, 64.1%) were 6.6 ±4.14 years old at testing. Developmental delay in the domains speech (n=158, 75.7%) and motor skills (n=153, 73.1%) were the most common findings. Furthermore, 135 patients had intellectual disability (64.4%), and 83 patients had epilepsy (39.9%) (**Figure 1)**.

**Figure 1.**
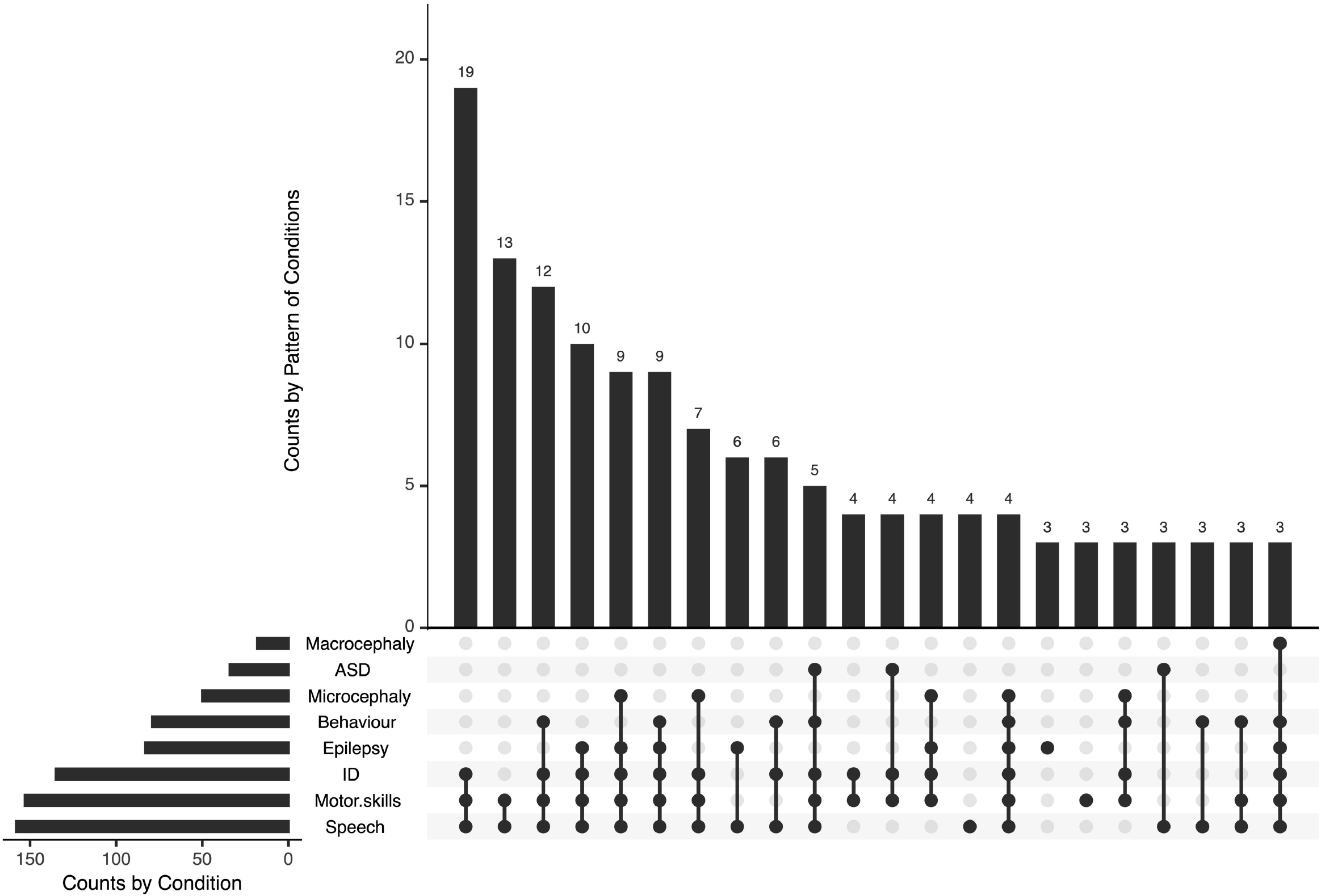
Phenotype of children of mothers tested for SYN1 autoantibodies. Developmental delay in the domains speech and motor skills were the most common findings, often associated with intellectual disability and epilepsy. Abbreviations: ID, intellectual disability; ASD, autism-spectrum disorder.

In a first step, the patients’ mothers were screened using the previously described CBA (14). We used a visual-scoring system for antibody binding in which scores of 2 were considered as positive (**Figure 2A**). Positive binding was identified in 20 mothers (9.6%). In 30 mothers, unspecific binding was detected while all others(n=158) showed no binding (**Figure 2B**). We further analyzed the sera of mothers with a SYN1 ELISA. Here, we found only a weak correlation between the CBA and ELISA results (r=0.29, **Figure 2C**). Possibly, these differing results can be explained by the binding of SYN1 autoantibodies to conformational epitopes that are not detected by ELISA. Therefore, we performed Western blots using the sera and cortex homogenates of wild-type and *SynI/II/III* triple knockout mice. We detected IgG binding to wild-type mouse brain homogenates in four of the 20 CBA-positive sera (**Figure 2D**). The other sera did not show immunoreactive bands at the expected molecular weight. These results point towards SYN1 IgG autoantibodies binding to linear SYN1 epitopes in only a small group, whereas autoantibodies against conformational synapsin epitopes predominate.

**Figure 2.**
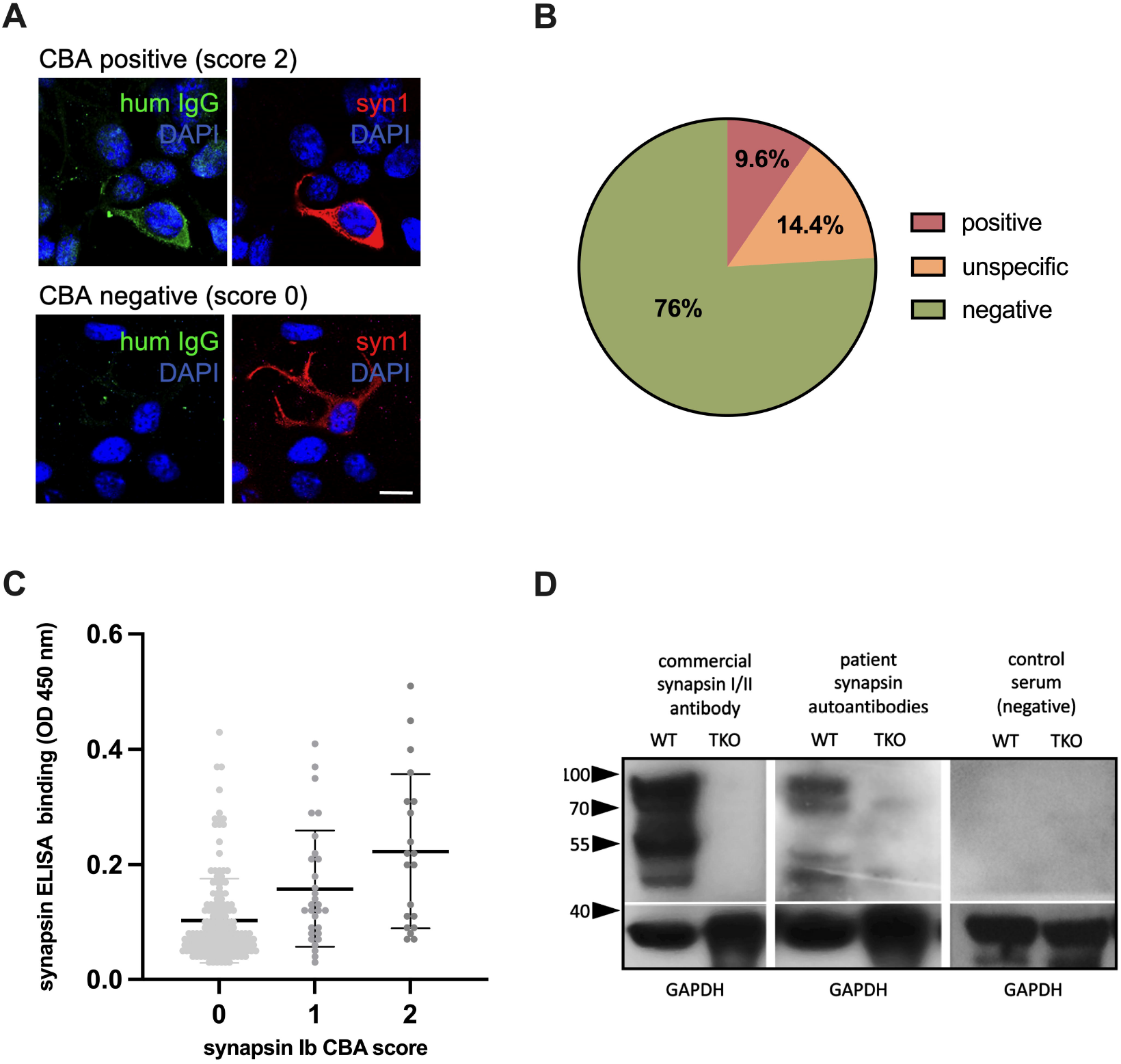
Detection of synapsin 1 (SYN1) autoantibodies. (**A**) Examples of immunofluorescence stainings, using human sera at a 1:300 dilution (green) with or without binding to HEK cells overexpressing human SYN1b are shown in the top row and bottom row. Using a commercial SYN1/2 antibody, protein expression is verified (red). DAPI is used to stain the nuclei (blue). Scale bar: 20 µm. (**B**) In 9.6% of mothers SYN1 autoantibodies were detected using a CBA. (**C**) CBA and in-house ELISA correlated weakly, with higher mean serum IgG levels in the CBA-positive group. (**D**) Representative immunoblots of wild type (WT) and Syn1/2/3 triple KO (TKO) mice cortex homogenates with a commercial synapsin 1/2 antibody as positive control, and with sera (1:200 dilution) from a CBA-positive mother and a CBA-negative control. Major bands at 80-90 kDa corresponding to the molecular weight of synapsin 1a/1b and additional bands at 50-55 kDa that could represent the synapsin 2b isoform or breakdown products of synapsin 1 were detected in wild type but not in TKO mouse tissue by both the commercial antibody and the serum of the CBA-positive mother. Detection of GAPDH served as loading control. Abbreviations: HEK human embryonic kidney; CBA, cell-based assay; ELISA, enzyme-linked immunosorbent assay, WB, western blot.

**Figure 3.**
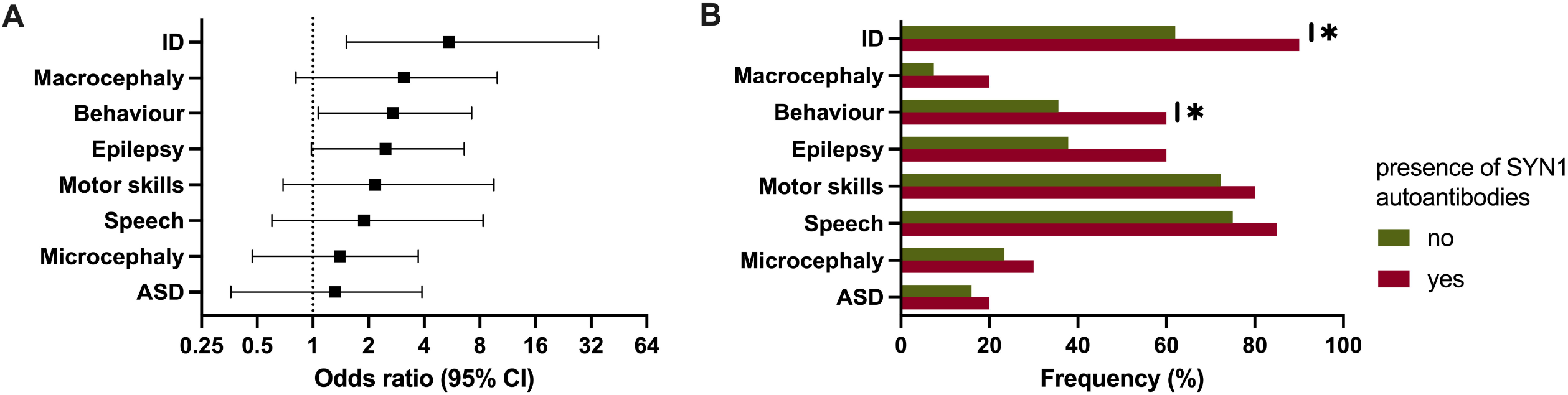
Clinical phenotype in patients of mothers with autoantibodies against synapsin 1 (SYN1). (**A, B**) Forest plot and bar chart showing the significantly increased risk for intellectual disability and behavioral problems. Abbreviations: ID, intellectual disability; ASD, autism-spectrum disorder.

We further analyzed children of mothers with a positive CBA (**Table 1)**. Most commonly the children had ID (n=18, 90%), speech delay (n=17, 85%), motor delay (n=17, 85%), and epilepsy (n=12, 60%). Seven patients had a developmental and epileptic encephalopathy (DEE). Cranial MRI was performed in 15 patients, with abnormalities detected in eight patients (53.3%, **Table 1**). Genetic testing was carried out in 13 patients, with variants (n=7) and microdeletions (n=1) identified in eight patients (61.5%). Of these seven variants, one was classified as pathogenic (14.3%), three as probably pathogenic (42.9%), and three as variants of unclear significance (42.9%). The four patients with IgG binding to wild-type mouse brain, showed no striking phenotypic differences to patients without binding (**Table 1)**.

**Table 1.** Phenotype of patients with positive maternal synapsin I (SYN1) autoantibodies. Abbreviations: ASD; autism-spectrum disorder; CM, Chiari malformation; DEE; Developmental and epileptic encephalopathy; DS, Dravet syndrome; HC; Hydrocephalus communicans; ID, intellectual disability; MAC, macrocephaly; MIC, microcephaly; MDLS, Miller-Dieker-Lissencephaly syndrome; NAD, no abnormality detected; n.p.; not performed; PCH, Pontocerebellar hypoplasia; SBH, subcortical band heterotopia; Genetic analysis: 1, chromosome analysis; 2, microarray-based comparative genomic hybridization (Array-CGH); 3, trio whole-exome analysis.

| sex | age at test<br>(range, years) | ID | cMRI | Epilepsy;<br>seizures | Gene<br>name | cDNA<br>position | ACMG | NM<br>number | OMIM | Speech | Motor<br>skills | ASD | Behavior | MIC | MAC |
| --- | --- | --- | --- | --- | --- | --- | --- | --- | --- | --- | --- | --- | --- | --- | --- |
| m | 6-10 | + | CM | DEE;<br>focal-onset atonic<br>seizures | *AIFM1* | c.1465G>A | III | 004208.3 | 300169 | + | + | - | + | - | + |
| m | 0-5 | + | HC | DEE;<br>generalized tonic-<br>clonic seizures | 1,2,3 |  |  |  |  | + | + | - | + | - | + |
| m | 0-5 | + | n.p. | - | 1,2 |  |  |  |  | + | + | + | + | - | - |
| m | 0-5 | + | PCH | DEE;<br>epileptic spasms | n.p. |  |  |  |  | + | + | - | + | + | - |
| m | 0-5 | - | peritrigonal,<br>localized<br>dysmyelination | - | *PTEN* | c.441A>T | IV | 000314 | 601728 | + | + | - | - | - | + |
| f | 0-5 | + | lissencephaly | DEE;<br>focal onset clonic<br>seizures without<br>awareness | microdel.<br>17p13.3 |  |  |  | 247200,<br>MDLS | + | + | - | - | - | - |
| m | 0-5 | + | delayed<br>myelination | DEE;<br>epileptic spasms | 1,2,3 |  |  |  | 617057 | + | + | + | - | + | - |
| m | 0-5 | + | NAD | focal onset<br>seizures without<br>awareness | n.p. |  |  |  |  | + | + | - | + | - | - |
| f | 11-15 | + | NAD | generalized tonic-<br>clonic seizures | 1,2 |  |  |  |  | + | - | - | + | - | - |
| m | 11-15 | + | NAD | DEE;<br>focal onset<br>seizures without<br>awareness | n.p. |  |  |  |  | + | + | - | + | + | - |
| m | 6-10 | + | NAD | - | *DPYD* | c.623G>T | III | 000110 | 612779 | + | + | + | + | - | - |
| m | 0-5 | + | NAD | - | n.p. |  |  |  |  | - | + | + | - | - | - |
| f | 11-15 | + | n.p. | - | *ANO5* | c.172C>T | IV | 213599.2 | 608662 | + | + | - | + | - | + |
| f | 16-20 | + | infra- and<br>supratentorial<br>atrophy | DEE;<br>focal onset<br>myoclonic seizures<br>without awareness | *SCN1A* | c.3887T>C | III | 001165963 | 182389,<br>DS | + | + | - | - | + | - |
| m | 0-5 | + | n.p. | - | *DDB1* | c.637G>A | IV | 001923 | 600045 | + | + | - | - | - | - |
| m | 0-5 | + | n.p. | - | n.p. |  |  |  |  | + | + | - | + | - | - |
| f | 0-5 | + | SBH | focal onset<br>myoclonic seizures<br>without awareness | DCX | c.814C>T | V | 000555 | 300121 | + | + | - | - | + | - |
| m | 0-5 | + | n.p. | - | 1 |  |  |  |  | + | + | - | + | - | - |
| m | 16-20 | + | NAD | generalized tonic-<br>clonic seizures | n.p. |  |  |  |  | - | - | - | + | + | - |
| m | 6-10 | - | NAD | epilepsy of unknow<br>cause with tonic<br>clonic seizures | n.p. |  |  |  |  | - | - | - | - | - | - |

We next analyzed the clinical data based on the results of the CBA. While there was no association between CBA and age of mothers, age at birth or children’s age, we found an increased risk for specific phenotypes in patients of mothers with positive CBA. A positive CBA was a significant risk factor in their children for the presence of ID (OR: 5.46 [95% CI: 1.52 -35.01], p = .0134) and behavioral problems (2.71 [1.07 -7.23], p = .0328). Furthermore, patients showed a tendency for the presence of macrocephaly (3.11 [0.81 -9.92], p = .0785), epilepsy (2.47 [0.98 -6.59], p = .0536), ASD (1.32 [0.36 -3.89], p = .7493), microcephaly (1.40 [0.47 -3.72], p = .5882), and a developmental delay in the domains speech (1.89 [0.60 -8.34], p = .4167) and motor skills (2.17 [0.69 -9.55], p = .2223) (**Figure 2A** and **B**).

## Discussion

In this study, we recruited a clinical cohort of 208 mothers and their children and analyzed the prevalence of maternal SYN1 autoantibodies and their association with developmental phenotypes. SYN1 IgG autoantibodies were detected through CBA in 9.6% of mothers with neurologically sick children. Most of the SYN1 CBA positive cases were negative on Western blots under denaturing conditions, suggesting a conformational dependency of their target binding, as similarly observed in other human anti-neuronal autoantibodies (18). In the study by Höltje et al. in a cohort of patients with various psychiatric and neurological disorders and SYN1 autoantibodies, binding was detected in immunoblots in about half of all patients (14). It is unclear whether in less denaturing conditions more SYN1 CBA positive cases would be positive. As in this study, we found IgG binding in immunoblots of 263 pregnant women screened for synapsin I autoantibodies in only 22.9% and found no correlation between ELISA and CBA antibody levels (16).

SYN1 is member of the family of synapsins, encoded by the three genes *SYN1, SYN2*, and *SYN3* in humans. SYN1 regulates the trafficking of synaptic vessels between the readily releasable pool and reserve pool. Furthermore, Syn1 moderates neuronal development through neurite outgrowth and synaptogenesis (2). *Syn1* mutant mice have severe epilepsy with generalized seizures (19).

We recently reported on a cohort of 263 pregnant women with seropositivity for SYN1 autoantibodies in 13.3%, matching the prevalence in this cohort (16). Seropositivity was associated with abnormalities of fetal development including intrauterine growth retardation. In this study of 208 children with various neurologic disorders, a positive maternal CBA showed a strong association with ID and behavioral problems, while there was and a trend towards other abnormalities such as macrocephaly and epilepsy. This is intriguing given that patients with LoF variants in *SYN1* display a similar phenotype with epilepsy, ID, ASD, macrocephaly, and behavioral problems (3-5). The lack of a clear association to these phenotypes can possibly be explained by the low statistical power of the study, requiring larger cohorts. In addition, due to the monocentric design and subsequent selection bias, multicenter studies need to be conducted to increase power.

When further analyzing the 20 CBA-positive patients seven had identified genetic variants/microdeletions of which one was considered pathogenic and three as likely pathogenic variants. This raises speculation as to whether genetic alterations in brain proteins may lead to neoantigen formation and increased autoimmunity. It is unclear whether maternal autoantibodies against SYN1 have a direct pathogenic effect on fetal development or rather represent a clinical biomarker of neuronal cell damage. It has been shown that SYN1 autoantibodies can reach the cytoplasmic site through FcγII/III-mediated endocytosis and thereby replicating the molecular *SYN1* LoF phenotype *in vitro* (20). Other experimental studies have demonstrated the direct pathogenic effect of maternal antineuronal autoantibodies such as NMDAR, Caspr2, and aquaporin-4 antibodies (10-13). Further, Caspr2 autoantibodies present during pregnancy are associated with ID and developmental disorders (21). However, functional studies on the effects of SYN1 autoantibodies on neurodevelopment are lacking and should be performed at molecular resolution using patient-derived monoclonal antibodies (22).

Although this study has several limitations, due to the heterogeneity of the pediatric cohort, it indicates a potential influence of maternal SYN1 autoantibodies on neurodevelopment. Further prospective studies are needed in which pregnant women are tested for maternal autoantibodies and their child’s development is closely monitored. Although these results do not currently trigger any direct therapeutic or diagnostic consequence, they could influence standard treatment of pregnant women in the long term and possibly lead to new therapeutic approaches.

## Data Availability

All data produced in the present study are available upon reasonable request to the authors.

## Acknowledgements

We thank Stefanie Bandura, Matthias Sillmann, Doreen Brandl, Antje Dräger, and Monika Majer for excellent technical assistance and study nurse support. J.K. and M.N. are participants in the Berlin Institute of Health (BIH)-Charité Junior Clinician Scientist Program. This work was supported by grants from the German Research Foundation (DFG) (grants FOR3004, PR1274/4-1, PR1274/5-1, PR1274/9-1), by the Helmholtz Association (HIL-A03), the German Federal Ministry of Education and Research (Connect-Generate 01GM1908D), the Einstein Stiftung Fellowship through the Günter Endres Fond, and the Sonnenfeld-Stiftung.

## Author Contributions

Conceptualization: A.K. and H.P.; Patient recruitment: I.B., K.M., T.U. and E.S., CBA and ELISA testing: I.B., J.K., H.F.R., M.H., D.M. and J.T; Western blotting: M.H.; Data collection, analysis, and visualization: I.B. and K.L.M.; Resources: A.K. and H.P.; Writing – original draft: I.B., K.L.M. Writing – review & editing: all authors.

## References

1. Hilfiker S, Benfenati F, Doussau F, Nairn AC, Czernik AJ, Augustine GJ, et al. Structural Domains Involved in the Regulation of Transmitter Release by Synapsins. J Neurosci (2005) 25(10):2658–69. doi: 10.1523/jneurosci.4278-04.2005.

2. Valtorta F, Benfenati F, Greengard P. Structure and Function of the Synapsins. J Biol Chem (1992) 267(11):7195–8.

3. Garcia CC, Blair HJ, Seager M, Coulthard A, Tennant S, Buddles M, et al. Identification of a Mutation in Synapsin I, a Synaptic Vesicle Protein, in a Family with Epilepsy. J Med Genet (2004) 41(3):183–6. doi: 10.1136/jmg.2003.013680.

4. Fassio A, Patry L, Congia S, Onofri F, Piton A, Gauthier J, et al. Syn1 Loss-of-Function Mutations in Autism and Partial Epilepsy Cause Impaired Synaptic Function. Hum Mol Genet (2011) 20(12):2297–307. Epub 20110325. doi: 10.1093/hmg/ddr122.

5. Giannandrea M, Guarnieri FC, Gehring NH, Monzani E, Benfenati F, Kulozik AE, et al. Nonsense-Mediated Mrna Decay and Loss-of-Function of the Protein Underlie the X-Linked Epilepsy Associated with the W356× Mutation in Synapsin I. PLoS One (2013) 8(6):e67724. Epub 20130620. doi: 10.1371/journal.pone.0067724.

6. Kreye J, Wright SK, van Casteren A, Stöffler L, Machule ML, Reincke SM, et al. Encephalitis Patient-Derived Monoclonal Gabaa Receptor Antibodies Cause Epileptic Seizures. J Exp Med (2021) 218(11). Epub 20210921. doi: 10.1084/jem.20210012.

7. Noviello CM, Kreye J, Teng J, Prüss H, Hibbs RE. Structural Mechanisms of Gaba(a) Receptor Autoimmune Encephalitis. Cell (2022) 185(14):2469-77.e13. doi: 10.1016/j.cell.2022.06.025.

8. Maljevic S, Møller RS, Reid CA, Pérez-Palma E, Lal D, May P, et al. Spectrum of Gabaa Receptor Variants in Epilepsy. Curr Opin Neurol (2019) 32(2):183–90. doi: 10.1097/wco.0000000000000657.

9. Han VX, Patel S, Jones HF, Dale RC. Maternal Immune Activation and Neuroinflammation in Human Neurodevelopmental Disorders. Nature Reviews Neurology (2021) 17(9):564–79. doi: 10.1038/s41582-021-00530-8.

10. García-Serra A, Radosevic M, Pupak A, Brito V, Ríos J, Aguilar E, et al. Placental Transfer of Nmdar Antibodies Causes Reversible Alterations in Mice. Neurol Neuroimmunol Neuroinflamm (2021) 8(1). Epub 20201110. doi: 10.1212/nxi.0000000000000915.

11. Mader S, Brimberg L, Vo A, Strohl JJ, Crawford JM, Bonnin A, et al. In Utero Exposure to Maternal Anti-Aquaporin-4 Antibodies Alters Brain Vasculature and Neural Dynamics in Male Mouse Offspring. Sci Transl Med (2022) 14(641):eabe9726. Epub 20220420. doi: 10.1126/scitranslmed.abe9726.

12. Brimberg L, Mader S, Jeganathan V, Berlin R, Coleman TR, Gregersen PK, et al. Caspr2-Reactive Antibody Cloned from a Mother of an Asd Child Mediates an Asd-Like Phenotype in Mice. Mol Psychiatry (2016) 21(12):1663–71. Epub 20161004. doi: 10.1038/mp.2016.165.

13. Jurek B, Chayka M, Kreye J, Lang K, Kraus L, Fidzinski P, et al. Human Gestational N-Methyl-D-Aspartate Receptor Autoantibodies Impair Neonatal Murine Brain Function. Ann Neurol (2019) 86(5):656–70. Epub 20190918. doi: 10.1002/ana.25552.

14. Höltje M, Mertens R, Schou MB, Saether SG, Kochova E, Jarius S, et al. Synapsin-Antibodies in Psychiatric and Neurological Disorders: Prevalence and Clinical Findings. Brain Behav Immun (2017) 66:125–34. Epub 20170718. doi: 10.1016/j.bbi.2017.07.011.

15. Piepgras J, Höltje M, Otto C, Harms H, Satapathy A, Cesca F, et al. Intrathecal Immunoglobulin a and G Antibodies to Synapsin in a Patient with Limbic Encephalitis. Neurology - Neuroimmunology Neuroinflammation (2015) 2(6):e169. doi: 10.1212/nxi.0000000000000169.

16. Bünger I, Kreye J, Makridis K, Höltje M, Rasmussen HF, van Hoof S, et al. Synapsin Autoantibodies During Pregnancy Are Associated with Fetal Abnormalities. medRxiv (2022):2022.09.23.22280284. doi: 10.1101/2022.09.23.22280284.

17. Milovanovic D, Wu Y, Bian X, De Camilli P. A Liquid Phase of Synapsin and Lipid Vesicles. Science (2018) 361(6402):604–7.

18. Prüss H. Autoantibodies in Neurological Disease. Nature Reviews Immunology (2021) 21(12):798–813. doi: 10.1038/s41577-021-00543-w.

19. Rosahl TW, Spillane D, Missler M, Herz J, Selig DK, Wolff JR, et al. Essential Functions of Synapsins I and Ii in Synaptic Vesicle Regulation. Nature (1995) 375(6531):488–93. doi: 10.1038/375488a0.

20. Rocchi A, Sacchetti S, De Fusco A, Giovedi S, Parisi B, Cesca F, et al. Autoantibodies to Synapsin I Sequestrate Synapsin I and Alter Synaptic Function. Cell Death Dis (2019) 10(11):864. Epub 20191114. doi: 10.1038/s41419-019-2106-z.

21. Coutinho E, Jacobson L, Pedersen MG, Benros ME, Nørgaard-Pedersen B, Mortensen PB, et al. Caspr2 Autoantibodies Are Raised During Pregnancy in Mothers of Children with Mental Retardation and Disorders of Psychological Development but Not Autism. J Neurol Neurosurg Psychiatry (2017) 88(9):718–21. Epub 20170601. doi: 10.1136/jnnp-2016-315251.

22. Duong SL, Prüss H. Molecular Disease Mechanisms of Human Antineuronal Monoclonal Autoantibodies. Trends Mol Med (2022). Epub 20221021. doi: 10.1016/j.molmed.2022.09.011.

